# Intercontinental and Regional Disparities in Cancer Burden: A Comprehensive Analysis of Trends and Projections Using 1990-2021 GBD Data with Forecasting to 2035

**DOI:** 10.1101/2024.11.03.24316671

**Authors:** Jinghao Liang, Yijian Lin, Hengrui Liang, Jihao Qi, Jingchun Ni, Hongmiao Lin, Jianxing He

## Abstract

**Background:** Understanding the intercontinental and regional cancer burden attributable to modifiable risk factors is crucial for developing effective prevention strategies. Using GBD 2021 data, this study aims to identify disparities in cancer burden, predict future trends across different continents, and inform targeted interventions.

**Methods:** This study utilized the GBD 2021 framework to comprehensively assess the cancer burden across four world regions, with a focus on age-standardized incidence rates (ASIR), age-standardized mortality rates (ASMR), and disability-adjusted life years (DALYs). We systematically analyzed cancer-related risk factors using associations extracted from the GBD database. Through the application of average annual percent change (AAPC) and the Bayesian age-period-cohort (BAPC) model, we forecasted the cancer burden in each region from 2022 to 2035.

**Findings:** In 2021, cancer incidence and mortality rates varied widely across continents. The Americas led with an incidence of 1,633.49 per 100,000 and a mortality rate of 116.097, while Europe showed 950.248 and 132.578, respectively. Asia reported 636.893 incidence and 112.329 mortality, and Africa recorded the lowest rates at 332.175 and 98.594. High mortality is associated with elevated DALYs, notably in Europe with 3,284.53 DALYs per 100,000, reflecting the highest cancer burden. In high-income regions, lifestyle-related cancers-linked to smoking, colorectal, and breast cancers-prevail, while low-income areas, particularly sub-Saharan Africa, experience infection-driven cancers like cervical cancer. Younger females under 35 bear a higher cancer burden than males, but after 70, the trend reverses significantly, with men having higher mortality across all ages. Smoking is the leading mortality risk in the Americas, Europe, and Asia, while unsafe sex dominates in Africa, correlating with high cervical cancer incidence. From 1990 to 2021, high BMI and blood glucose have surged as cancer risk factors, driven by obesity and metabolic diseases. By 2035, projections indicate incidence will increase to 1,779.6 per 100,000 in the Americas, with stable mortality at 98.97, while Asia, Africa, and Europe are expected to see moderate incidence and mortality shifts, reflecting regional disparities in cancer prevention and management strategies.

**Interpretation:** This study reveals pronounced global disparities in cancer burden, shaped by economic development, healthcare access, and lifestyle factors. High-income regions, like North America and Europe, show high incidence but lower mortality rates due to robust screening and preventive measures. Conversely, low-income regions, especially sub-Saharan Africa, experience high mortality from preventable cancers, highlighting critical healthcare gaps. Gender analysis shows women have higher incidence due to screening, while men face higher mortality risks. Projected increases in incidence globally underscore the urgency for tailored cancer control strategies, focusing on prevention and healthcare access improvements across diverse socio-economic contexts.

## INTRODUCTION

Cancer is the second leading cause of death worldwide, imposing a significant burden on public health and socio-economic systems. According to the International Agency for Research on Cancer (IARC), it is estimated that in 2022, there were nearly 20 million new cancer cases worldwide, with approximately 9.7 million cancer-related deaths^1^. In recent years, with the aging global population, rapid urbanization, and changes in lifestyle, cancer incidence and mortality have shown a marked upward trend^2–5^. The burden of cancer not only has profound implications for individual health but also exerts substantial pressure on healthcare systems and impedes socio-economic development^6^. Despite the overall increase in the global cancer burden, significant disparities exist between different continents and regions, reflecting variations in socio-economic status, cultural background, access to healthcare services, and environmental factors^7–9^. For instance, in economically developed regions such as Europe and North America, cancer incidence rates are relatively high, but due to widespread screening, increased health awareness, and abundant healthcare resources, mortality rates are comparatively lower. In contrast, in Africa and parts of Asia, insufficient screening, limited healthcare resources, and challenges in accessing health services result in significantly higher cancer mortality^7^. Understanding these disparities is crucial for developing targeted control strategies, as different regions need to adopt interventions tailored to their specific needs to mitigate the cancer burden.

In 2015, the United Nations introduced the Sustainable Development Goals (SDGs), with Goal 3.4 aiming to reduce premature mortality from non-communicable diseases, including cancer, by one-third by 2030^10^. However, the COVID-19 pandemic has severely disrupted cancer screening and treatment services in many regions, leading to setbacks in the progress of cancer control. The pandemic not only interrupted routine cancer prevention and treatment services but also caused delays in patient consultations and reduced early cancer diagnosis rates, further exacerbating the cancer burden^11,12^. Effectively addressing these challenges requires the promotion of comprehensive interventions, including primary prevention, screening, and early treatment, tailored to the specific circumstances of different continents and regions. In particular, in economically underdeveloped areas, strengthening healthcare infrastructure, raising public health awareness, and ensuring access to medications and treatment are key to reducing the cancer burden^13^.

The Global Burden of Disease (GBD) study provides a unique data resource that systematically assesses the contribution of various modifiable risk factors to the cancer burden^14–16^. The GBD study is currently among the most comprehensive resources, encompassing a broad range of countries, age groups, and genders, enabling systematic quantification of cancer burdens across various time points and regions. The most recent iteration, GBD 2021, presents an updated dataset encompassing the global burden of cancer across 204 countries and territories from 1990 to 2021^17–19^. This iteration introduces refined methodologies and updated insights, ensuring the database remains highly relevant to contemporary health priorities and emerging challenges.

This study, based on the latest data from GBD 2021, conducts an in-depth analysis of the cancer burden across the four major continents (Africa, the Americas, Asia, and Europe) and their respective regions. By quantitatively assessing cancer incidence and mortality across continents and regions, as well as their temporal dynamics, we aim to identify the major risk factors in each region and provide a scientific basis for the development of targeted public health interventions. This study will provide empirical support for the formulation of global and regional cancer control policies, helping to achieve the goal of reducing the cancer burden and offering practical intervention recommendations for different regions.

## Methods

### Overview

The Global Burden of Disease (GBD) study was founded to provide standardized, comprehensive global health estimates, encompassing mortality, disability, and related risk factors across all countries. The 2021 iteration of the GBD study offers extensive data on 371 diseases and injuries alongside 88 risk factors across 204 countries and territories, covering the period from 1990 to 2021^18,20^. All input data and statistical codes are publicly accessible through the Global Health Data Exchange (GHDx), and the GBD study rigorously adheres to the Guidelines for Accurate and Transparent Health Estimates Reporting (GATHER)(https://www.who.int/publications/m/item/gather-checklist), ensuring transparency and reproducibility of global health metrics (<u>Global Burden of Disease Study 2021 (GBD 2021) Code | GHDx</u>).

### Overview of Global Cancer Burden Estimates

Cancers included in the GBD study correspond to those defined in Chapter 2 (neoplasms) of the tenth revision of the International Classification of Diseases^14^. The study synthesizes data from a diverse array of high-quality sources, including population-based cancer registries, vital statistics systems, hospital and clinical records, peer-reviewed literature, and the GBD’s international collaborator network. In instances where direct incidence data is unavailable, mortality-to-incidence ratios (MIRs) are employed, adjusted through the Healthcare Access and Quality Index to identify and mitigate potential outliers. Mortality estimation leverages data from cancer registries, official vital records, and verbal autopsy data. In cases of sparse mortality data, incidence records are converted into mortality estimates via MIRs, tailored to specific cancer types^15,21^. Additionally, the Cause of Death Ensemble Modeling (CODEm) framework is utilized to assess individual and ensemble models, refining predictions with a focus on optimal predictive performance across regions, age groups, and years^22^. Final mortality estimates are aligned with independently modeled all-cause mortality data to maintain consistency across demographic and temporal parameters^9^. Non-fatal cancer burden is captured through YLDs and DALYs, with the model tracking the cancer disease course through four phases—diagnosis, remission, metastatic spread, and terminal stages—each adjusted with sequela-specific disability weights, thereby producing YLDs^14^. Meanwhile, YLLs are derived by applying age-specific mortality data to the remaining standard life expectancy at each age^23^. Ultimately, DALYs were derived as the aggregate of YLDs and YLLs.

### Risk Factors

In the GBD study framework, risk factors are systematically organized within a four-tier hierarchical structure, commencing with a comprehensive grouping of all risks. At Level 1, risk factors are categorized into three primary domains: environmental and occupational, behavioral, and metabolic. At Level 2, these domains are further delineated into 20 specific clusters, such as dietary risks and air pollution. Progressing to Level 3, nine of these clusters are subdivided into 42 distinct risk factors. Finally, Level 4 provides a detailed breakdown of five Level 3 risk factors into 22 additional sub-categories^16^. Building upon the GBD’s core metrics—namely, mortality, and DALYs—this study aims to quantify the impact of specific risk factors on the burden of all neoplasms. Our primary focus is on secondary-level risk factors. For each of these, we calculated population attributable fractions (PAFs) by integrating exposure data, relative risk estimates, and theoretical minimum risk exposure levels (TMRELs). Stratified by location, age, sex, and year, the PAFs provide estimates of the potential reduction in neoplasm burden if exposures were minimized to optimal levels. To translate these PAFs into measurable health outcomes, each PAF was multiplied by the DALYs, thereby quantifying the burden attributable to each risk factor both regionally and globally^16^. Our analysis encompasses global estimates as well as those for four major world regions: Asia, Africa, the Americas, and Europe. By structuring our approach in this manner, we facilitate a comparative analysis of how modifiable risk factors contribute to neoplasm burdens across diverse socio-demographic contexts. This comprehensive assessment not only highlights the global impact of neoplasms but also identifies modifiable risk factors as strategic targets for region-specific prevention efforts.

### Statistical Analysis

We acquired data on annual incidence, mortality, DALYs, and age-standardized rates (ASRs) for neoplasms and 33 subtypes of neoplasms (Appendix Table 1) from the Global Health Data Exchange (https://vizhub.healthdata.org/gbd-results/). We conducted a comprehensive analysis of cancer statistics on a global scale, focusing specifically on four world regions: the Americas, Africa, Asia, and Europe, six World Health Organization (WHO) regions: the African Region, the Region of the Americas, the South-East Asia Region, the Eastern Mediterranean Region, the Western Pacific Region, and the European Region and seven World Bank regions: Europe & Central Asia, East Asia & Pacific, Latin America & Caribbean, Sub-Saharan Africa, North America, the Middle East & North Africa, and South Asia. Emphasizing cancer-related risk factors, we focus on estimates for all neoplasms and a subset of 23 specific cancers within the GBD dataset (Appendix Table 1). We present absolute, proportional, and age-standardized rates (ASRs) of incidence (ASIR) and mortality (ASMR) for cancer at global, regional, and national levels^17^. Key trends in cancer burden are highlighted through comparisons across age groups, sexes, and geographic regions. Temporal changes in ASRs were assessed via the average annual percent change (AAPC), calculated using Joinpoint Trend Analysis Software (Version 5.2.0), which provides a weighted summary of annual percent changes (APCs). Statistically significant trends (determined by two-sided tests) are classified as “increasing” or “decreasing,” while non-significant trends are labeled “stable^24^.” Future trend predictions leveraged the Integrated Nested Laplace Approximations (INLA) framework with the Bayesian age-period-cohort (BAPC) model, effectively addressing challenges of mixing and convergence associated with traditional Bayesian methods^25^. Based on GBD data from 1990 to 2021 and population projections from the WHO, the BAPC model accurately forecasts future trends. All statistical analyses and visualizations were conducted using R (version 4.4.1), with additional R packages including “ggplot2,” “RColorBrewer,” “patchwork,” and “ggrepel.” All hypothesis tests were conducted as two-tailed, with statistical significance defined at P < 0.05.

## RESULTS

In 2021, cancer DALYs, incidence, and mortality rates varied significantly across continents (Figure1 A-C). In Asia, Mongolia had the highest DALYs, exceeding 5,000 per 100,000, while most South and East Asian countries ranged between 1,500 and 3,000. In Europe, Eastern countries like Romania and Poland recorded DALYs above 3,000, while Western European nations were generally below 2,500. In the Americas, Greenland approached 4,000 DALYs, while most of the United States and South American countries ranged from 2,000 to 3,000. In Africa, South Africa and Zimbabwe reported the highest DALYs, exceeding 3,500, with most other African countries below 2,000 (Figure1 A). Mortality rates were highest in Asia’s Mongolia and Africa’s South Africa, both exceeding 200 per 100,000. In Europe, Eastern countries showed rates around 150, while Western Europe and most of South America had lower rates, below 100(Figure1 B). Incidence rates were highest in North America, with parts of the United States reaching nearly 4,000 per 100,000, followed by Western Europe, where rates approached 3,000. Most Asian countries reported incidence rates below 1,500, and Africa had the lowest rates, with most countries under 1,000(Figure1 C). Overall, DALYs and mortality rates showed strong alignment, with higher DALYs often associated with higher mortality. Notable examples included Asia’s Mongolia and Africa’s South Africa and Zimbabwe, while Western Europe and parts of the northern United States had lower levels in both metrics(Figure1 A-C).

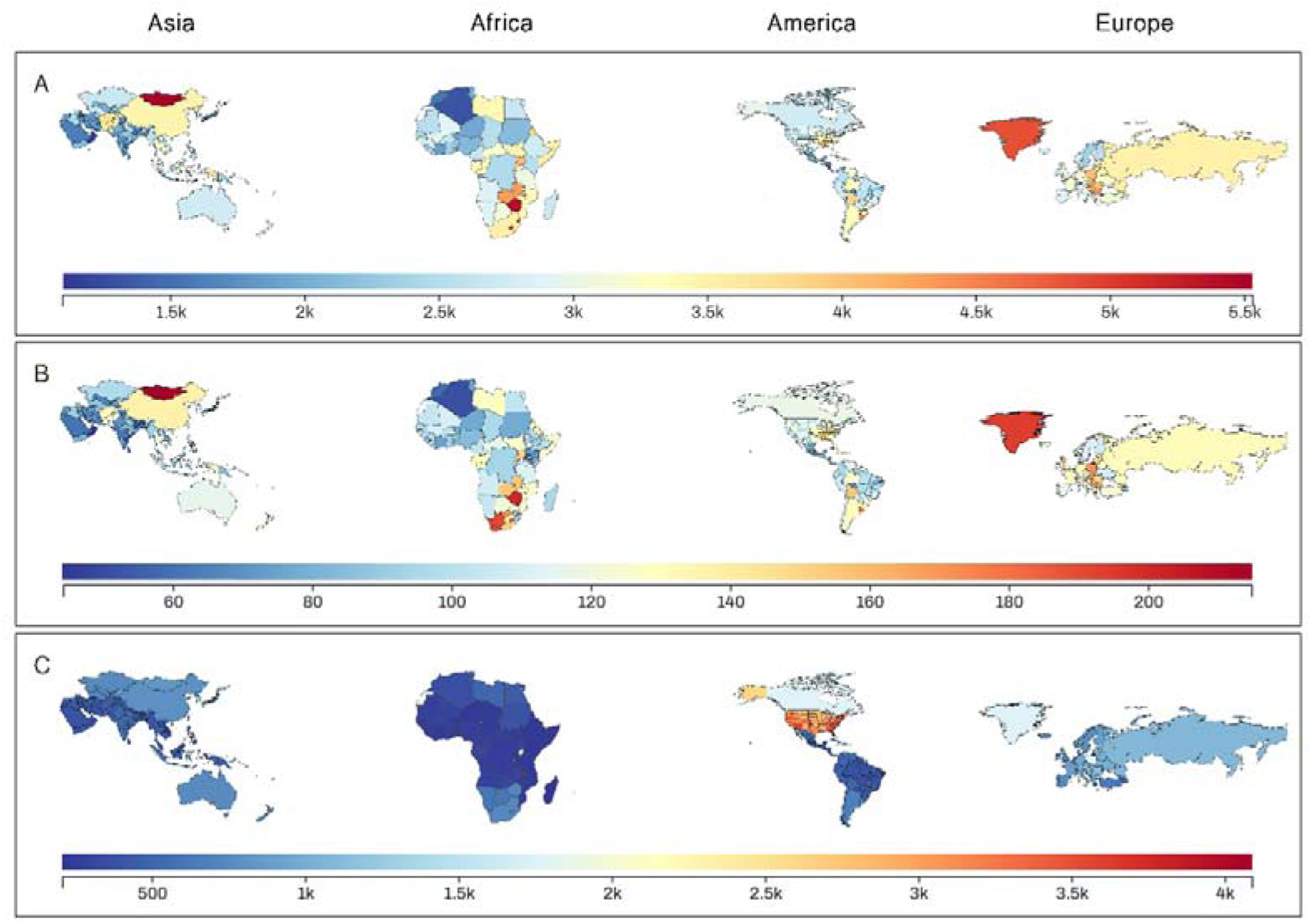
P1 : National cancer burden across the four world regions for 2021. The maps illustrate age-standardized rates of (C) incidence, (A) disability-adjusted life years (DALYs), and (B) mortality for each country within the four world regions.

In 2021, cancer burden varied significantly across continents in terms of DALYs, mortality, and incidence (Figure2 A-B, Table1). In Africa, bladder cancer imposed the highest DALY burden at 2,614.9 per 100,000 people, followed by eye cancer (310.9) and gallbladder cancer (263.9). Similarly, in the Americas, bladder cancer had the highest DALY impact (2,946.6), alongside multiple myeloma (472.6) and nasopharyngeal cancer (275.8). In Asia, cervical cancer contributed most to the DALY burden (557.3), followed by Hodgkin lymphoma (262.3) and stomach cancer (133.6). In Europe, the largest DALY burdens were from soft tissue cancers (657.5), uterine cancer (376.4), and stomach cancer (285.5) (Table 1). Mortality rates also reflected regional variations, with bladder cancer leading cancer-related deaths in Africa (2.5 per 100,000), followed by cervical cancer (8.2) and tracheal, bronchus, and lung cancer (8.9). In the Americas, tracheal, bronchus, and lung cancer accounted for the highest mortality rate (21.5), followed by colorectal cancer (12.3) and breast cancer (9.3). In Asia, tracheal, bronchus, and lung cancer were the primary causes of cancer mortality (24.9), followed by stomach cancer (13.9) and breast cancer (6.1). In Europe, tracheal, bronchus, and lung cancer had the highest mortality rate (27.3), with colorectal cancer (16.9) and breast cancer (10.3) following (Figure2 A-B, E Table1). Incidence rates revealed further differences, with non-melanoma skin cancer driving the highest incidence in the Americas (357.6 per 100,000), followed by breast cancer (37.5) and prostate cancer (35.0). Europe’s incidence was primarily due to colorectal cancer (37.3) and breast cancer (37.7), with a total rate of 950.2. In Asia, tracheal, bronchus, and lung cancer (27.7) and stomach cancer (18.3) were the main contributors to an incidence rate of 636.9. Africa had the lowest incidence overall, largely due to cervical cancer (13.0) and breast cancer (18.6), totaling 332.2 per 100,000 (Figure2 A-B, E Table1).

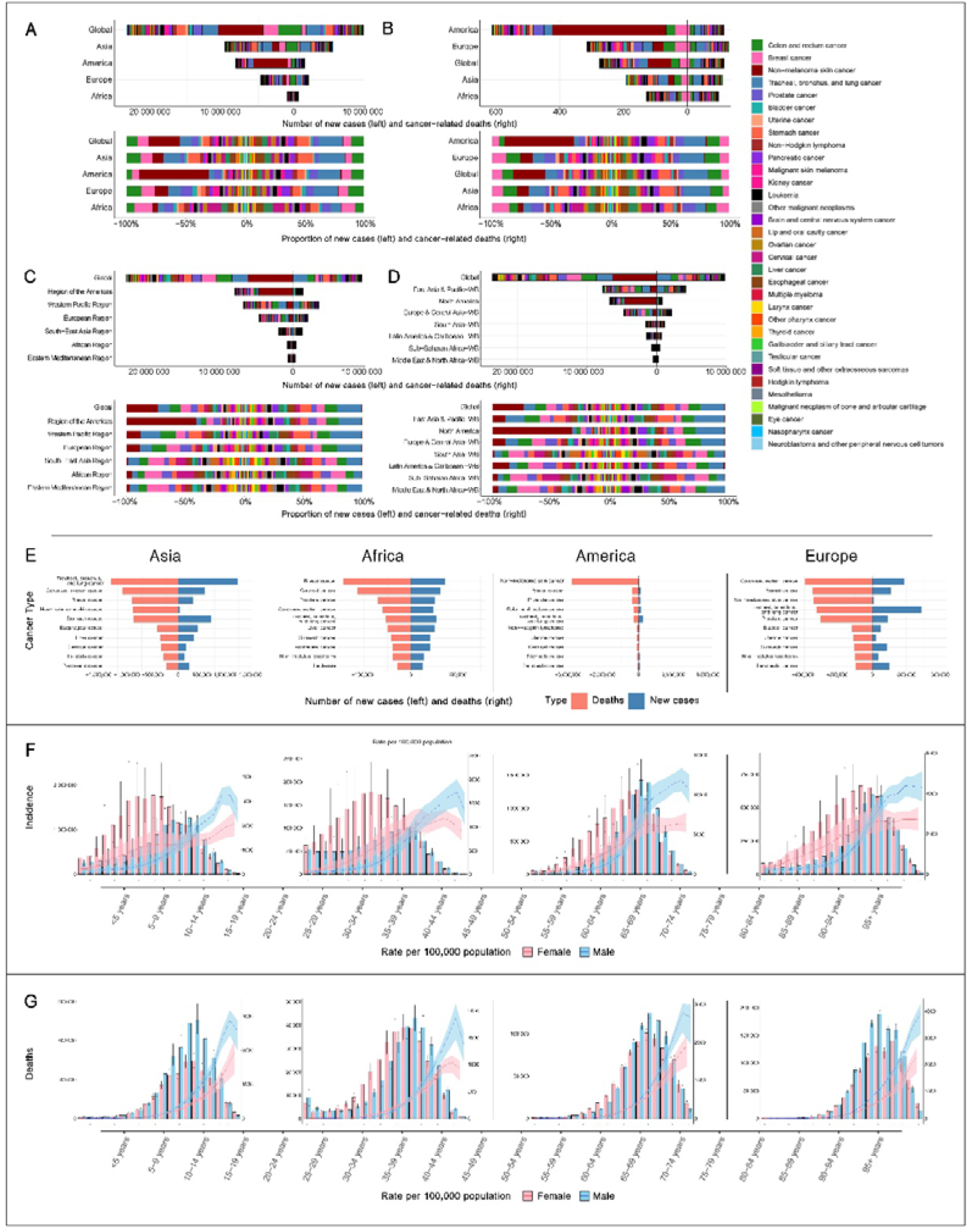
P2 : Global and regional cancer burden for 2021. (F-G) Age-specific counts and rates of incident cases and deaths by sex across the four world regions. (E) Counts of incident cases and deaths for the ten most common cancers in the four world regions. (A-B) Counts, rates, and proportions of incident cases and deaths categorized by cancer type across the four world regions. (C) Counts and proportions of incident cases and deaths by cancer type within World Health Organization (WHO) regions. (D) Counts and proportions of incident cases and deaths by cancer type within World Bank regions.

An analysis based on World Bank economic stratification indicates that in high-income regions (such as North America and Europe), the cancer burden is predominantly associated with lifestyle-related cancers, including lung cancer, colorectal cancer, and breast cancer (Figure2 D). In North America and Europe, lung cancer accounts for 24.9% and 20.4% of mortality, respectively. Colorectal cancer’s incidence rate is 3.6% in North America and 12.6% in Europe, while breast cancer has an incidence rate of 4.4% in North America and accounts for 7.7% of cancer mortality in Europe. In contrast, in low-income regions (such as Sub-Saharan Africa), the cancer burden is significantly driven by infection-related factors. Cervical cancer in these regions has an incidence rate of 14.6% and a mortality rate of 11.3%, being one of the leading causes of cancer mortality among women. Liver cancer accounts for 7.0% of cancer-related deaths. When categorized by the World Health Organization’s regional divisions, the distribution of cancers aligns closely with regional culture and lifestyle (Figure2 C). For instance, the Americas have a very high incidence of non-melanoma skin cancer associated with excessive ultraviolet radiation exposure, at 357.6 cases per 100,000 people. Southeast Asia exhibits a high incidence of lip and oral cavity cancer, related to chewing habits, at 7.5 cases per 100,000 people. In the Western Pacific, stomach cancer, associated with preserved food consumption, and liver cancer, associated with hepatitis B infection, have high incidence rates of 26.8 and 10.2 per 100,000 people, respectively. From a gender perspective, cancer burden trends by gender are similar across regions: from under 5 years old to around 70 years, the cancer burden in females surpasses that in males; after this age, the trend reverses, with males bearing a higher burden than females. With aging, both the absolute numbers and rates of incidence and mortality rise markedly in both sexes, with the increase being particularly rapid in males (Figure2 F-G).

In 2021, the global cancer burden exhibited significant regional disparities across the four continents. In Northern Asia, Mongolia showed markedly high disability-adjusted life years (DALYs) (5,530 per 100,000 people) and cancer mortality (215 per 100,000 people), with Central and East Asia also presenting notably high values, reflecting an extremely high disease burden. Similarly, Greenland in Europe ranks high in DALYs (4,862.2 per 100,000) and mortality (193.9 per 100,000), indicating relatively poor health conditions in the region, while most Eastern and Western European countries exhibit moderate levels, with Eastern European countries such as Hungary and Poland showing slightly higher cancer burdens than Western Europe. In the Americas, there is a notable north-south disparity, with a higher burden in South America, especially in parts of Chile and Argentina, where DALYs in Chile approach 4,000, while North America, particularly Canada, has a comparatively lower cancer mortality rate within the Americas. Africa presents the most diverse situation, with the highest burden observed in Southern Africa. Zimbabwe shows a particularly high cancer mortality rate (201.3 per 100,000 people), significantly higher than other African nations, while Southern African countries like South Africa and Namibia also show elevated burdens. In contrast, North Africa has a lower burden, with countries such as Morocco (mortality rate of 58.2 per 100,000) experiencing relatively low cancer burdens within the continent (figure1 A).

The ranking of cancer risk factors across four continents reveals several data-driven trends. In 2021, smoking remained the leading cancer mortality risk in the Americas, Europe, and Asia, consistent with its 1st or 2nd position in 1990, underscoring its persistent carcinogenic impact in these regions. In contrast, unsafe sex ranked as the top cancer risk factor in Africa from 1990 through 2021, highlighting a regional distinction likely associated with the high prevalence of cervical cancer and HPV infection in this area. High body mass index rose in rank across all continents, placing 2nd or 3rd in the Americas, Europe, and Africa in 2021, and advancing from 15th to 7th in Asia from 1990 to 2021, reflecting the increasing global impact of obesity on cancer mortality. Additionally, high fasting plasma glucose climbed in rank on all continents, reaching 3rd in the Americas, 5th in Europe, 4th in Asia, and 6th in Africa in 2021, indicating the growing role of metabolic diseases, such as diabetes, in cancer burden worldwide. Particulate matter exposure consistently ranked among the top 10 cancer risk factors across all continents in 2021, particularly in Asia and Africa, where it ranked 2nd and 5th, respectively. Occupational carcinogen exposure (e.g., asbestos and benzene) held prominent positions in 2021, ranking 4th in the Americas and 3rd in Europe, which correlates with higher levels of industrialization in these regions. The sustained prominence of smoking in the Americas, Europe, and Asia, alongside the rise of obesity and metabolic risk factors across all continents, reflects a global trend toward lifestyle-related cancer risk, while distinct regional factors, such as unsafe sex in Africa and high occupational carcinogen exposure in industrialized regions, illustrate the diverse public health challenges in cancer prevention globally (Figure3).

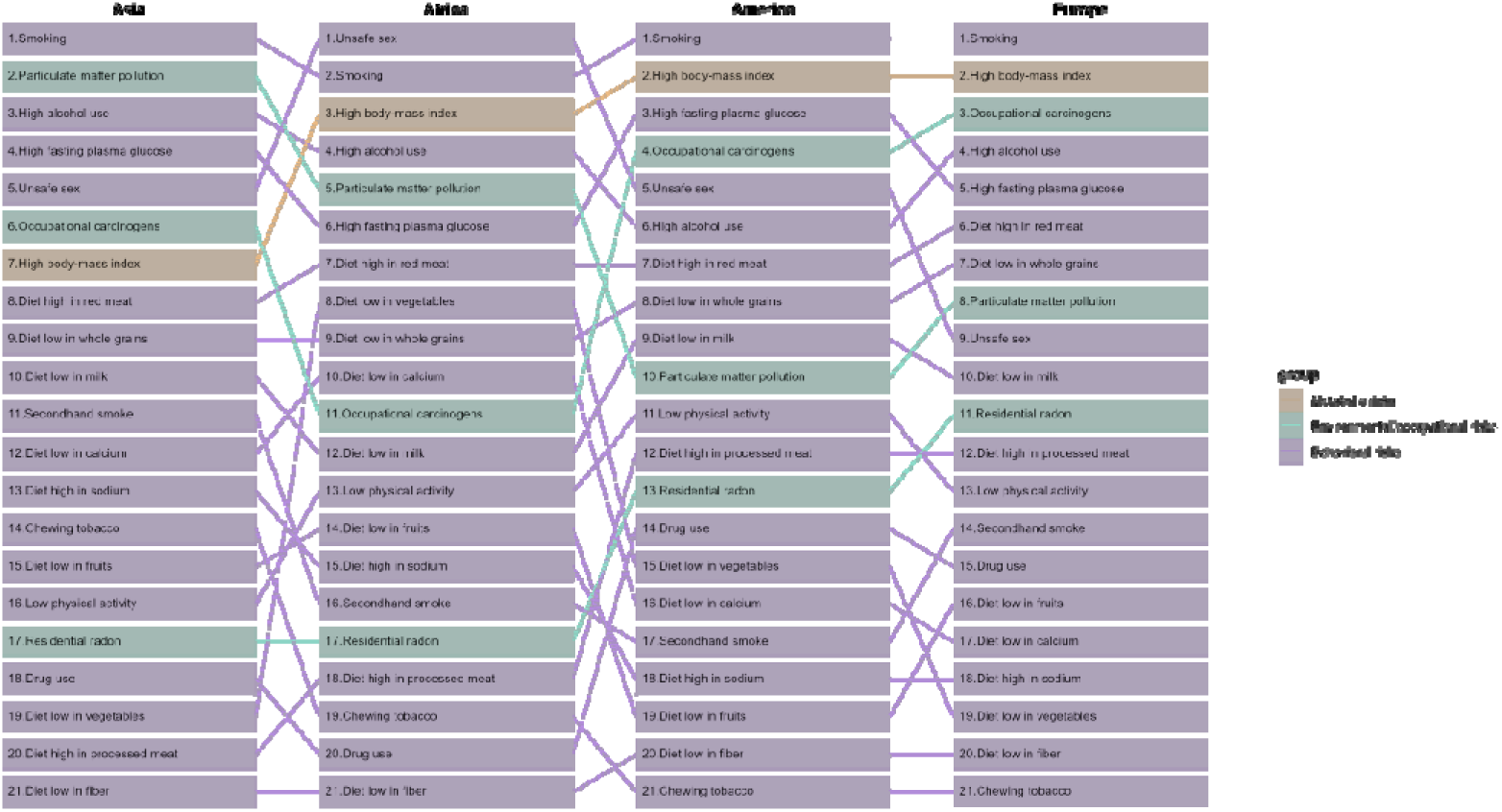
P3 : The ranking of cancer risk factors across four continents in 2021. These risk factors are represented by different colors, reflecting their types (orange represents metabolic risks, green represents environmental and occupational risks, and purple represents behavioral risks).

From 1990 to 2021, the trend in cancer mortality rates across the globe and the four continents shows clear regional disparities and cancer-specific characteristics (Figure4 A-B). Globally, mortality rates for most major cancers have significantly declined, especially for stomach, esophageal, and cervical cancers, with annual average percentage changes (AAPCs) of : −2.17%, −1.19%, and −1.25%, respectively, reflecting global progress in preventing and controlling these cancers. However, regional trends for specific cancers vary. In Africa, breast and pancreatic cancer mortality rates have significantly increased, with AAPCs of 1.1% and 1.47%, respectively, while stomach and cervical cancer mortality rates have declined by −0.85% and −0.6% AAPC, respectively, highlighting an increased demand for breast and pancreatic cancer prevention and control. The Americas generally show a decrease in mortality rates, with breast cancer (−1.1% AAPC) and lung cancer (−1.69% AAPC) exhibiting particularly pronounced declines, possibly due to strengthened early screening and risk factor control. Asia shows a marked decline in mortality rates for stomach and esophageal cancers, with AAPCs of −2.26% and −1.66%, suggesting effective targeted preventive measures in this region; however, breast cancer mortality has increased by 0.57% AAPC, indicating a need to further strengthen prevention strategies. Europe has seen the greatest decline in stomach cancer mortality (−2.89% AAPC), reflecting significant progress in controlling this cancer; however, pancreatic cancer mortality has slightly increased (0.35% AAPC), indicating its continued high lethality. Overall, global and continental cancer mortality rates show a general downward trend, but distinct regional differences are evident for specific cancers (Figure4 C-D).

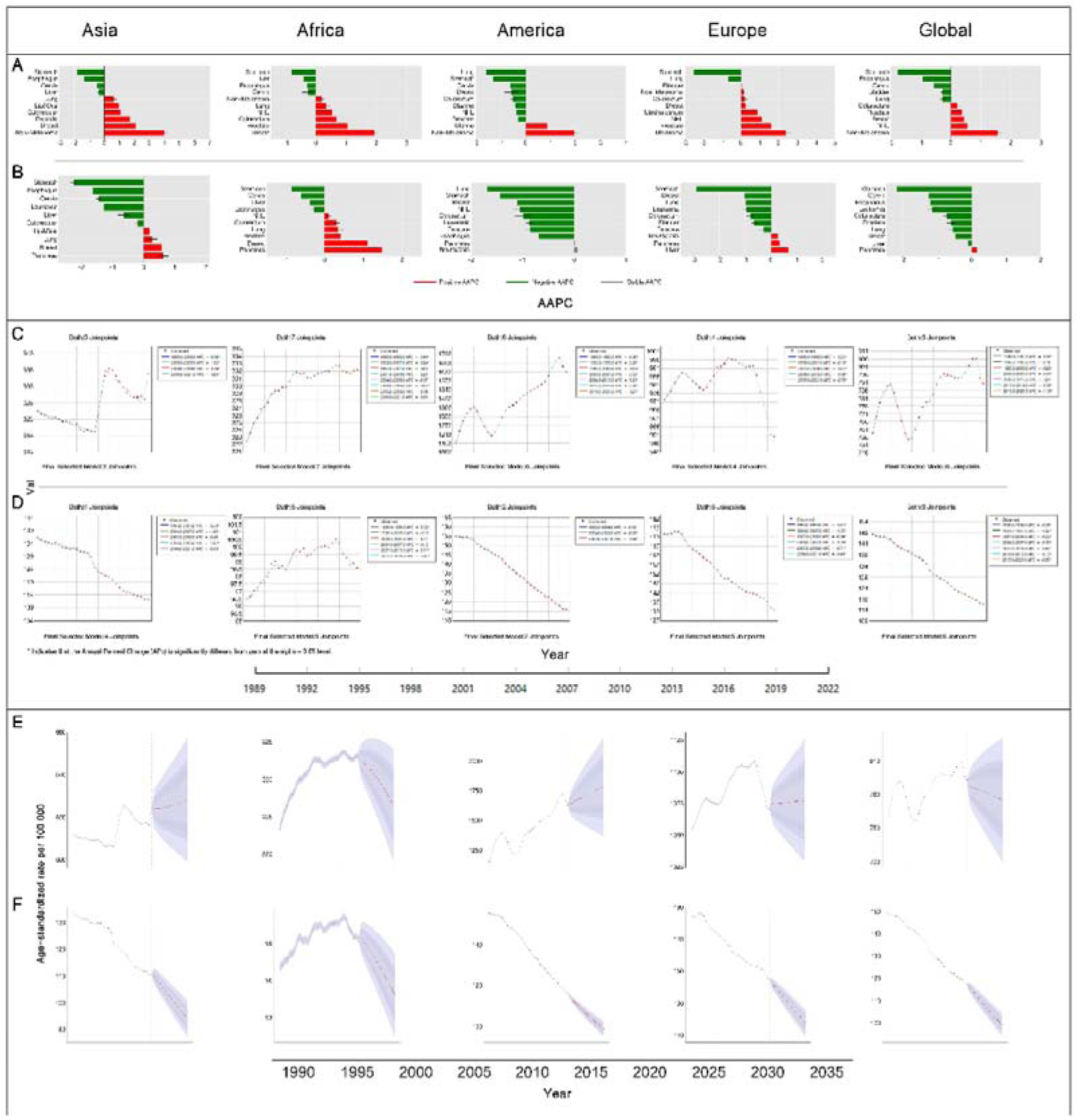
P4: Temporal trends in the cancer burden globally and across the four world regions. (A-B) Trends for the ten most frequent cancer types from 1990 to 2021. (C-D) Trends by year from 1990 to 2021. (E-F) Projections from 2022 to 2035 using the BAPC model. Abbreviations: AAPC, average annual percent change; BAPC, Bayesian age-period-cohort. *Indicates statistical significance at P < 0.05.

From 2022 to 2035, projected data indicate distinct trends in cancer incidence and mortality globally and across the continents. In the Americas, the incidence rate is projected to increase significantly from 1,640.4 per 100,000 people in 2022 to 1,779.6 per 100,000 people in 2035, while the mortality rate is expected to remain relatively stable, with a projected 2035 value of approximately 98.97 per 100,000 people. In Asia, the incidence rate is projected to increase slightly from 623.9 per 100,000 people in 2022 to 628.1 per 100,000 people in 2035, with a slight decrease in mortality to an estimated 94.21 per 100,000 people in 2035. In Africa, both incidence and mortality rates are projected to see small increases, with the incidence rate rising from 332.5 per 100,000 people to 326.5 per 100,000 people, and mortality expected to rise to approximately 94.84 per 100,000 people during the same period. Europe’s incidence rate is projected to remain relatively unchanged, increasing slightly from 1,074.4 per 100,000 people in 2022 to 1,077.4 per 100,000 people in 2035, while the mortality rate is expected to continue decreasing, reaching 117.79 per 100,000 people by 2035. Globally, the incidence rate is expected to gradually rise to 774.8 per 100,000 people by 2035, while the mortality rate is projected to decrease slightly, with a predicted value of 99.22 per 100,000 people(Figure4 E-F).

## DISCUSSION

This study reveals marked disparities in cancer incidence and mortality rates across different global regions, closely tied to each region’s healthcare resources, policy implementation, and cultural background. Data indicate that high-income regions, such as North America and Europe, exhibit higher cancer incidence, particularly for lifestyle-related cancers like colorectal and lung cancer, which are strongly associated with high-fat diets, physical inactivity, and smoking behaviors^26^. These regions have broadly implemented smoking cessation policies and early screening programs, contributing to relatively low cancer mortality rates.^27–29^ However, the cumulative long-term effects of smoking remain evident in the high incidence of lung cancer^30^. In contrast, low-income regions, such as Africa and Southeast Asia, continue to bear high mortality rates for preventable cancers like cervical and breast cancer due to limited medical resources. Sub-Saharan Africa, in particular, has the highest global burden of cervical cancer, attributed to low HPV vaccination rates and inadequate screening coverage^31^. High-income countries have significantly reduced cervical cancer incidence through vaccination and screening initiatives, whereas low-income areas face a rising burden of preventable cancers due to inadequate policy coverage^32–34^. Cultural practices also exert significant influence on cancer prevalence patterns. For example, the high incidence of oral cancer in Southeast Asia is closely associated with betel nut chewing^35^, while the elevated rates of stomach and liver cancer in the Western Pacific region are linked to high-salt diets, preserved foods, and hepatitis B infection^36–38^. Therefore, tailored interventions are needed: HPV vaccination and cervical cancer screening programs should be expanded in low-income countries, and culturally sensitive health education is necessary in Southeast Asia and the Western Pacific to reduce betel nut and preserved food consumption, aiming to alleviate the cancer burden in these regions.

Notably, our findings highlight significant gender differences in global cancer data: while women generally exhibit higher cancer incidence than men, men experience significantly higher cancer mortality. This discrepancy is likely a result of multiple factors, including biological differences, health behaviors, and socioeconomic conditions. Women are more likely to participate in routine screenings for cancers such as breast and cervical cancer, which increases the likelihood of early detection and thus raises overall incidence rates in women^39,40^. Additionally, estrogen levels are strongly associated with increased susceptibility to breast and endometrial cancers, especially in postmenopausal women, where the risk becomes more pronounced^41^. In men over 50, higher testosterone levels correlate with an increased risk of aggressive prostate cancers^42–44^. On the other hand, men tend to have higher rates of smoking^27^, alcohol consumption, and occupational exposures to high-risk environments (such as chemical and mining industries), which elevate their risk for cancers with high fatality rates^45^. Men are also more likely to delay medical consultation, leading to late-stage diagnoses, limited treatment options, and lower survival rates. Socioeconomic factors further exacerbate the gender disparities in cancer burden. In high-income populations, better medical resources allow women greater access to cancer screenings and early treatment^46^, while men face higher cancer mortality risks due to occupational exposures and reluctance to seek timely medical care. In low-income and resource-constrained environments, both genders have limited access to healthcare, yet men’s higher occupational exposure increases their cancer mortality risk substantially^47^. Overall, gender disparities in cancer incidence and mortality reflect multi-layered inequalities, underscoring the need for targeted cancer prevention, screening measures, and increased awareness of early diagnosis in men to reduce their elevated cancer mortality risk.

This study demonstrates that, between 1990 and 2021, dynamic shifts in cancer risk factors across continents strongly reflect the impact of economic development, urbanization, and cultural and lifestyle differences on cancer burden. In regions experiencing rapid industrialization and urbanization, environmental carcinogens from air pollution and occupational exposures have significantly increased, particularly in Asia, where particulate pollution continues to exacerbate respiratory cancer burdens—a risk intensified by industrial and urban expansion, highlighting the trade-offs between economic growth and environmental risks^48^. In economically advanced regions, cancer burden is more closely associated with metabolic and lifestyle-related factors. For instance, in North America and Europe, the rising consumption of red and processed meats, alongside increases in obesity and hyperglycemia, has significantly raised the risks of colorectal and liver cancers^47,49,50^. While these regions have widely implemented anti-smoking policies^28^, the long-term cumulative effects of smoking remain evident in lung cancer prevalence, reflecting the lag between policy implementation and historical exposure^27^. In contrast, cancer risks in economically developing countries are primarily driven by infectious and occupational factors. In these regions, limited healthcare resources, insufficient vaccine coverage, and low screening rates contribute to high incidences of cancers associated with HIV and other infectious diseases^51^. Additionally, the lack of effective occupational protections has led to prolonged exposure to carcinogens, further increasing cancer risk among workers^52–54^. These regional disparities underscore the critical need for targeted policies. In rapidly industrializing areas, stricter pollution controls and the promotion of clean energy are essential. In North America and Europe, where the burden of metabolic diseases and smoking remains high, rigorous dietary management and obesity prevention policies should be prioritized. Meanwhile, in countries with lower levels of economic development, resources should be directed toward vaccination programs, infectious disease prevention, and occupational safety to effectively reduce cancer burdens^47^.

The projections for the global and regional cancer burden from 2022 to 2035 highlight significant disparities across continents. In the Americas, incidence rates are expected to rise while mortality rates remain relatively stable, suggesting that although therapeutic advances have been made, persistent risk factors continue to drive incidence, underscoring the need for enhanced prevention and early screening^55^. Asia shows moderate growth in incidence; however, given its large population, the overall cancer burden is projected to increase substantially, reflecting varying responses to urbanization and pollution-associated health risks. Africa faces a dual rise in both incidence and mortality, underscoring severe deficiencies in healthcare resources and the urgent need to improve cancer care infrastructure. In Europe, relatively stable incidence rates alongside declining mortality are attributed to comprehensive screening and treatment approaches, though aging populations may impose new challenges on healthcare systems. Globally, while mortality shows a downward trend, the rising incidence accentuates the need for widespread cancer prevention efforts, particularly in low-resource settings. Reducing risk factors, expanding vaccination coverage, and bolstering cancer care capacity in underserved regions are essential for achieving long-term control over the cancer burden. These forecasts emphasize the urgency of promoting health equity and developing region-specific public health policies to address the global cancer challenge.

This study, consistent with other GBD analyses, faces inherent limitations due to the unique modeling approaches and data sources employed by GBD, as previously documented^16–18^. One key limitation pertains to regional classification: in GBD, Oceania is grouped with the Asia region, which may obscure significant geographic, cultural, and healthcare system differences. Oceania and Asia differ substantially in health risk profiles, resource availability, and disease burden, and this regional aggregation may mask health issues specific to Oceania, thereby diminishing the precision and applicability of regional policy recommendations. Future studies should aim to conduct separate analyses for these regions to better capture their distinct health needs. Another significant limitation concerns the variability in data quality and availability, particularly from low- and middle-income regions. Sparse epidemiological data from these regions may introduce biases, thereby affecting the reliability of both mortality and incidence estimates. In areas where primary data are limited, GBD relies on extrapolation and modeled estimates, which, despite their methodological rigor, remain inherently constrained by gaps in data coverage and inconsistencies in diagnostic and reporting practices. These limitations underscore the need for enhanced data infrastructure and improved quality of cancer surveillance globally. The impact of the COVID-19 pandemic also introduces a degree of uncertainty to the mortality and morbidity estimates presented, especially in regions severely affected by the pandemic. The data utilized in this analysis are from 2021, during the ongoing pandemic, and thus do not reflect post-pandemic conditions. Assessing the interaction between COVID-19 and cancer outcomes remains challenging, as the pandemic likely influenced cancer diagnosis rates, treatment availability, and patient health-seeking behaviors—factors which complicate projections of the long-term cancer burden. Furthermore, the emphasis on regional disparities in this study may have inadvertently resulted in a lack of granular analysis at the country level. This lack of country-specific focus may obscure unique national patterns of cancer burden, limiting the ability to offer precise, targeted public health interventions for individual nations with specific health challenges. Lastly, while GBD’s approach to risk factor analysis is comprehensive, it may not capture all relevant risk factors for cancer across regions. Emerging or less-studied risk factors, particularly those unique to localized populations, may be inadequately represented. Additionally, reliance on existing literature may overlook novel or localized exposures that are not yet well characterized.

## Data Availability

All data generated in this study are available upon reasonable request from the authors. All data generated in this study are included in the manuscript. All data generated are accessible online.

https://www.healthdata.org/research-analysis/gbd

## References

1. Bray, F., et al. Global cancer statistics 2022: GLOBOCAN estimates of incidence and mortality worldwide for 36 cancers in 185 countries. CA. Cancer J. Clin. 74, 229–263 (2024).

2. van Hoogstraten, L. M. C. et al. Global trends in the epidemiology of bladder cancer: challenges for public health and clinical practice. Nat. Rev. Clin. Oncol. 20, 287–304 (2023).

3. Ramaboli, M. C. et al. Diet changes due to urbanization in South Africa are linked to microbiome and metabolome signatures of Westernization and colorectal cancer. Nat. Commun. 15, 3379 (2024).

4. Zare Sakhvidi, M. J., et al. Exposure to greenspace and cancer incidence, prevalence, and mortality: A systematic review and meta-analyses. Sci. Total Environ. 838, 156180 (2022).

5. Domínguez-Berjón, M. F., Gandarillas, A. & Soto, M. J. Lung cancer and urbanization level in a region of Southern Europe: influence of socio-economic and environmental factors. J. Public Health 38, 229–236 (2016).

6. Chen, S. et al. Estimates and Projections of the Global Economic Cost of 29 Cancers in 204 Countries and Territories From 2020 to 2050. JAMA Oncol. 9, 465 (2023).

7. Sung, H., et al. Global Cancer Statistics 2020: GLOBOCAN Estimates of Incidence and Mortality Worldwide for 36 Cancers in 185 Countries. CA. Cancer J. Clin. 71, 209–249 (2021).

8. Roth, G. A. et al. Global, regional, and national age-sex-specific mortality for 282 causes of death in 195 countries and territories, 1980–2017: a systematic analysis for the Global Burden of Disease Study 2017. The Lancet 392, 1736–1788 (2018).

9. Vos, T. et al. Global burden of 369 diseases and injuries in 204 countries and territories, 1990–2019: a systematic analysis for the Global Burden of Disease Study 2019. The Lancet 396, 1204–1222 (2020).

10. Transforming our world: the 2030 Agenda for Sustainable Development | Department of Economic and Social Affairs. https://sdgs.un.org/2030agenda.

11. Wang, H. et al. Estimating excess mortality due to the COVID-19 pandemic: a systematic analysis of COVID-19-related mortality, 2020–21. The Lancet 399, 1513–1536 (2022).

12. Sharpless, N. E. COVID-19 and cancer. Science 368, 1290–1290 (2020).

13. Farmer, P. et al. Expansion of cancer care and control in countries of low and middle income: a call to action. The Lancet 376, 1186–1193 (2010).

14. Tran, K. B. et al. The global burden of cancer attributable to risk factors, 2010–19: a systematic analysis for the Global Burden of Disease Study 2019. The Lancet 400, 563–591 (2022).

15. Murray, C. J. L. et al. Global burden of 87 risk factors in 204 countries and territories, 1990–2019: a systematic analysis for the Global Burden of Disease Study 2019. The Lancet 396, 1223–1249 (2020).

16. Brauer, M. et al. Global burden and strength of evidence for 88 risk factors in 204 countries and 811 subnational locations, 1990–2021: a systematic analysis for the Global Burden of Disease Study 2021. The Lancet 403, 2162–2203 (2024).

17. Ferrari, A. J. et al. Global incidence, prevalence, years lived with disability (YLDs), disability-adjusted life-years (DALYs), and healthy life expectancy (HALE) for 371 diseases and injuries in 204 countries and territories and 811 subnational locations, 1990–2021: a systematic analysis for the Global Burden of Disease Study 2021. The Lancet 403, 2133–2161 (2024).

18. Naghavi, M. et al. Global burden of 288 causes of death and life expectancy decomposition in 204 countries and territories and 811 subnational locations, 1990–2021: a systematic analysis for the Global Burden of Disease Study 2021. The Lancet 403, 2100–2132 (2024).

19. Schumacher, A. E. et al. Global age-sex-specific mortality, life expectancy, and population estimates in 204 countries and territories and 811 subnational locations, 1950–2021, and the impact of the COVID-19 pandemic: a comprehensive demographic analysis for the Global Burden of Disease Study 2021. The Lancet 403, 1989–2056 (2024).

20. Murray, C. J. L. Findings from the Global Burden of Disease Study 2021. The Lancet 403, 2259–2262 (2024).

21. Global Burden of Disease 2019 Cancer Collaboration. Cancer Incidence, Mortality, Years of Life Lost, Years Lived With Disability, and Disability-Adjusted Life Years for 29 Cancer Groups From 2010 to 2019: A Systematic Analysis for the Global Burden of Disease Study 2019. JAMA Oncol. 8, 420–444 (2022).

22. Foreman, K. J., Lozano, R., Lopez, A. D. & Murray, C. J. Modeling causes of death: an integrated approach using CODEm. Popul. Health Metr. 10, 1 (2012).

23. Wang, H. et al. Global age-sex-specific fertility, mortality, healthy life expectancy (HALE), and population estimates in 204 countries and territories, 1950–2019: a comprehensive demographic analysis for the Global Burden of Disease Study 2019. The Lancet 396, 1160–1203 (2020).

24. Li, W. et al. Global cancer statistics for adolescents and young adults: population based study. J. Hematol. Oncol.J Hematol Oncol 17, 99 (2024).

25. Knoll, M. et al. An R package for an integrated evaluation of statistical approaches to cancer incidence projection. BMC Med. Res. Methodol. 20, 257 (2020).

26. Banks, J., Marmot, M., Oldfield, Z. & Smith, J. P. Disease and Disadvantage in the United States and in England. JAMA 295, 2037–2045 (2006).

27. Groot, P. M. de, Wu, C. C., Carter, B. W. & Munden, R. F. The epidemiology of lung cancer. Transl. Lung Cancer Res. 7, (2018).

28. Bryazka, D. et al. Forecasting the effects of smoking prevalence scenarios on years of life lost and life expectancy from 2022 to 2050: a systematic analysis for the Global Burden of Disease Study 2021. Lancet Public Health 9, e729–e744 (2024).

29. Qaseem, A. et al. Screening for Colorectal Cancer in Asymptomatic Average-Risk Adults: A Guidance Statement From the American College of Physicians. Ann. Intern. Med. 171, 643–654 (2019).

30. Leiter, A., Veluswamy, R. R. & Wisnivesky, J. P. The global burden of lung cancer: current status and future trends. Nat. Rev. Clin. Oncol. 20, 624–639 (2023).

31. Forman, D. et al. Global Burden of Human Papillomavirus and Related Diseases. Vaccine 30, F12–F23 (2012).

32. Arbyn, M. et al. Estimates of incidence and mortality of cervical cancer in 2018: a worldwide analysis. Lancet Glob. Health 8, e191–e203 (2020).

33. Brisson, M. et al. Impact of HPV vaccination and cervical screening on cervical cancer elimination: a comparative modelling analysis in 78 low-income and lower-middle-income countries. The Lancet 395, 575–590 (2020).

34. Catarino, R., Petignat, P., Dongui, G. & Vassilakos, P. Cervical cancer screening in developing countries at a crossroad: Emerging technologies and policy choices. World J. Clin. Oncol. 6, 281–290 (2015).

35. Zhang, X. & Reichart, P. A. A review of betel quid chewing, oral cancer and precancer in Mainland China. Oral Oncol. 43, 424–430 (2007).

36. Wogan, G. N., Hecht, S. S., Felton, J. S., Conney, A. H. & Loeb, L. A. Environmental and chemical carcinogenesis. Semin. Cancer Biol. 14, 473–486 (2004).

37. Shikata, K. et al. A prospective study of dietary salt intake and gastric cancer incidence in a defined Japanese population: The Hisayama study. Int. J. Cancer 119, 196–201 (2006).

38. Tsugane, S. & Sasazuki, S. Diet and the risk of gastric cancer: review of epidemiological evidence. Gastric Cancer 10, 75–83 (2007).

39. The benefits and harms of breast cancer screening: an independent review. The Lancet 380, 1778–1786 (2012).

40. Smith, R. A. et al. Cancer screening in the United States, 2019: A review of current American Cancer Society guidelines and current issues in cancer screening. CA. Cancer J. Clin. 69, 184–210 (2019).

41. Liang, Y. et al. Global burden and trends in pre- and post-menopausal gynecological cancer from 1990 to 2019, with Projections to 2040: a cross-sectional study. Int. J. Surg. 10.1097/JS9.0000000000001956 doi:10.1097/JS9.0000000000001956.

42. Snyder, P. J. et al. Effects of Testosterone Treatment in Older Men. N. Engl. J. Med. 374, 611–624 (2016).

43. Platz, E. A. & Giovannucci, E. The epidemiology of sex steroid hormones and their signaling and metabolic pathways in the etiology of prostate cancer. J. Steroid Biochem. Mol. Biol. 92, 237–253 (2004).

44. Morgentaler, A. & Traish, A. M. Shifting the Paradigm of Testosterone and Prostate Cancer: The Saturation Model and the Limits of Androgen-Dependent Growth. Eur. Urol. 55, 310–321 (2009).

45. Steenland, K. Laryngeal cancer incidence among workers exposed to acid mists (United States). Cancer Causes Control 8, 34–38 (1997).

46. Louwman, W. J., van de Poll-Franse, L. V., Fracheboud, J., Roukema, J. A. & Coebergh, J. W. W. Impact of a programme of mass mammography screening for breast cancer on socio-economic variation in survival: a population-based study. Breast Cancer Res. Treat. 105, 369–375 (2007).

47. Jemal, A., Center, M. M., DeSantis, C. & Ward, E. M. Global Patterns of Cancer Incidence and Mortality Rates and Trends. Cancer Epidemiol. Biomarkers Prev. 19, 1893–1907 (2010).

48. Lelieveld, J., Evans, J. S., Fnais, M., Giannadaki, D. & Pozzer, A. The contribution of outdoor air pollution sources to premature mortality on a global scale. Nature 525, 367–371 (2015).

49. Aune, D. et al. Red and processed meat intake and risk of colorectal adenomas: a systematic review and meta-analysis of epidemiological studies. Cancer Causes Control 24, 611–627 (2013).

50. Bosch, F. X., Ribes, J., Díaz, M. & Cléries, R. Primary liver cancer: Worldwide incidence and trends. Gastroenterology 127, S5–S16 (2004).

51. Rendle, K. A. et al. Patient perspectives on delays in cervical cancer screening and follow-up care in Botswana: a mixed methods study. BMC Womens Health 22, 195 (2022).

52. Richter, E. D. et al. Cancer risks in naval divers with multiple exposures to carcinogens. Environ. Health Perspect. 111, 609–617 (2003).

53. Naito, S. et al. Cancer occurrence among dyestuff workers exposed to aromatic amines. A long term follow-up study. Cancer 76, 1445–1452 (1995).

54. Jayaraj, S. & Shiva Nagendra, S. M. Health risk assessment of workers’ exposure to BTEX and PM during refueling in an urban fuel station. Environ. Monit. Assess. 195, 1507 (2023).

55. Humphrey, L. L. et al. Screening for Lung Cancer With Low-Dose Computed Tomography: A Systematic Review to Update the U.S. Preventive Services Task Force Recommendation. Ann. Intern. Med. 159, 411–420 (2013).

